# Nascent prostate cancer heterogeneity drives evolution and resistance to intense hormonal therapy

**DOI:** 10.1101/2020.09.29.20199711

**Authors:** Scott Wilkinson, Huihui Ye, Fatima Karzai, Stephanie A. Harmon, Nicholas T. Terrigino, David J. VanderWeele, John R. Bright, Rayann Atway, Shana Y. Trostel, Nicole V. Carrabba, Nichelle C. Whitlock, Stephanie M. Walker, Rosina T. Lis, Houssein A. Sater, Brian J. Capaldo, Ravi A. Madan, James L. Gulley, Guinevere Chun, Maria J. Merino, Peter A. Pinto, Daniela C. Salles, Harsimar B. Kaur, Tamara L. Lotan, David J. Venzon, Peter L. Choyke, Baris Turkbey, William L. Dahut, Adam G. Sowalsky

## Abstract

Localized prostate cancer is distinctively characterized by intratumoral heterogeneity, and tumors with more complex evolutionary paths display more aggressive characteristics. In clinical trials of intense neoadjuvant androgen deprivation therapy (inADT), patients with complete or near-complete responses experience durable remissions. However, the molecular characteristics distinguishing exceptional responders and nonresponders at baseline have not been established. Here, we present the integrated histologic and genomic analysis of pre-treatment baseline tissue from our recent Phase 2 clinical study of inADT. Multiregion sampling demonstrated that patients with incomplete and nonresponding tumors demonstrate greater tumor diversity as estimated by phylogenetic tree reconstruction from DNA sequencing and automated analysis of immunohistochemical stains. Development of a four-factor binary model to predict poor response correlated with increased genomic diversity in our 37-patient cohort and a validation cohort of 188 Gleason score 8-10 prostate cancers. Together, these findings demonstrate that even in the primary setting, more highly evolved tumors have increased fitness to resist therapy.

## INTRODUCTION

Although prostate cancer is the most common non-cutaneous malignancy in American males^1^, the vast majority of cases are detected and treated early when prostate cancer is potentially curable by surgery or radiation ^2^. In contrast, localized prostate tumors that progress despite definitive therapy are characterized by extensive evolutionary heterogeneity that manifests itself both at the time of initial treatment and later upon relapse, where therapy resistance in metastatic, castration-resistant prostate cancer (mCRPC) can be associated with numerous subclones that emerged from a common ancestor ^3,4^. Recently, numerous case series have identified subsets of human tumors, including prostate cancers, that are at increased risk of recurrence due to greater intratumoral heterogeneity, and evidence suggests that active mutational processes are more likely to give rise to a daughter clone with resistance in more evolutionarily advanced cancer systems ^5-7^. For patients with prostate cancer, who may go for more than a decade between initial therapy and development of radiographically confirmed metastases, intervening chemohormonal treatments and their effects on tumor progression are based on biochemical recurrence (rising serum PSA levels) with rare opportunities for sampling tumors by biopsy until later in the disease course. Consequently, little is known about the relationship between the evolutionary trajectory of treatment-naïve localized prostate tumors and their propensity to resist direct treatment.

The androgen receptor (AR) remains a molecular target for prostate cancer therapy, both by the inhibition of the release of testicular androgen (androgen deprivation therapy, or ADT) by gonadotropin hormone releasing hormone agonists and newer-generation inhibitors of dihydroxytestosterone biosynthesis such as abiraterone and direct AR-inhibition including enzalutamide, apalutamide and darolutamide. These agents have been approved by the United States Food and Drug Administration for treatment of mCRPC but recent clinical trials have demonstrated their efficacy in nonmetastatic CRPC (*i.e*. early ADT-resistant prostate cancer) as well as in *de novo* metastatic hormone sensitive prostate cancer ^8-11^. Although the rationale for these drugs’ activity earlier in the disease course is that AR activity is critical for disease progression, it is logical that inhibition of AR activity can ablate organ-confined disease or prevent it from spreading.

Neoadjuvant ADT, and more recently intense neoadjuvant ADT (*i.e*. ADT plus one or more AR-targeted agents), has been tested in a series of Phase 2 clinical trials in efforts to decrease intratumoral testosterone levels and diminish the volume of residual disease prior to surgery ^12-14^. Indeed, these studies have demonstrated that baseline tumor volume is a significant predictor of final pathology volumes, but have also indicated that patients with less residual disease at the end of treatment experience longer durations of biochemical recurrence free survival ^15^. With pathologic complete response rates variable across cohorts, molecular analyses of post-treatment surgical specimens have indicated that molecular alterations potentially driving resistance are subclonal and present prior to treatment, drawing potential relationships between the extent of a given tumor’s evolution and increased fitness associated with subclonal heterogeneity in a hormone sensitive state^16,17^.

We thus hypothesized that evolutionary principles relating diversity and heterogeneity to clinical outcome could explain in part why larger tumors at baseline could result in diverse resistance mechanisms post-treatment. In this study, we present the integrated histologic and genomic analysis of 141 laser capture microdissected tumor exomes representing 110 baseline biopsy tissues from 37 patients with intermediate-to-high risk prostate cancer who were enrolled in a phase 2, single-arm clinical trial of neoadjuvant androgen deprivation therapy intensified by the addition of enzalutamide ^18^. Using both tissue- and DNA-based analyses, we observe that the volume of residual disease at the end of therapy increases proportionally with tumoral diversity at baseline. Localized prostate cancers that are more evolved are thus more likely to be resistant to intense systemic hormonal therapy and have a lower chance of relapse-free survival. To predict the likelihood of non-response, we demonstrate that a straightforward four-factor histogenomic model recapitulates the relationship between non-response and increased baseline subclonal diversity. Our results demonstrate that increased fitness in more evolved tumors contributes, at least in part, to treatment resistance in high risk localized prostate cancer.

## METHODS AND MATERIALS

### Biospecimen procurement

This trial was approved by the Institutional Review Board of the Center for Cancer Research, National Cancer Institute (ClinicalTrials.gov identifier: NCT02430480). This study has been conducted in accordance with ethical principles that have their origin in the Declaration of Helsinki and are consistent with the International Council on Harmonization guidelines on Good Clinical Practice, all applicable laws and regulatory requirements, and all conditions required by a regulatory authority and/or institutional review board. Written informed consent was obtained for all 39 patients prior to performing study-related procedures in accordance with federal and institutional guidelines. Patients underwent a multiparametric MRI (mpMRI) upon enrollment and MRI/US-fusion biopsies were targeted to one or more lesions identified on mpMRI. In many cases, a 12-core templated biopsy was also performed. A sample of saliva (Oragene OG500, DNA Genotek) or blood, from which genomic DNA was further derived, was also procured prior to treatment initiation. Following six months of ADT (either goserelin, leuprolide or degarelix at the discretion of the treating physician) plus enzalutamide (160 mg administered orally, daily for 24 weeks), patients underwent a second mpMRI prior to radical prostatectomy. Two patients did not complete the study and are censored from analysis. One patient underwent transurethral resection of the prostate (TURP) due to extension of the prostate tumor to the bladder, and consequently final pathology volumes for that patient are censored despite his classification as a nonresponder to therapy.

### Treatment response

At the conclusion of therapy, all but three study subjects underwent radical prostatectomy (RP). The RP specimen was grossed and sectioned in the same plane as the mpMRI, using a customized 3-D printed mold designed using the most recent post-treatment mpMRI ^19^. Surgical specimens were stained with H&E and additional immunostains to verify the presence of residual tumor, including anti-NKX 3.1, PIN-4 cocktail, CAM5.2, and anti-p63 ^18,20^. Determination of residual disease was performed by three genitourinary pathologists (M.J.M., H.Y., and R.T.L.). Residual cancer burden (RCB) was calculated by multiplying the number of slices containing residual tumor by the largest cross-sectional width and length of that tissue and by block thickness (0.6 cm). Volume was further corrected by multiplying by 0.4 to account for tumor cellularity.

We considered RCB < 0.05 cc as exceptional responders (ER) and we grouped incomplete and nonresponders (INR) into a single category for RCB > 0.05 cc. Each mpMRI-visible lesion was similarly classified based on the volume evaluable tumor remaining in the anatomical region of the lesion. If a patient had more than one residual lesion at final pathology, the largest cross-sectional dimension of tumor was used to determine RCB for the purpose of classifying the patient as ER or INR. This approach resulted in more ER lesions than ER patients, as non-index responding lesions were harbored by INR patients.

### Biopsy selection

From 37 patients, 62 mpMRI-visible lesions were identified and sampled by biopsy (up to 4 lesions per patient). For larger lesions, additional sampling by biopsy of the lesion was performed to account for potential heterogeneity. Of the 62 sampled lesions, 9 did not have any tumor and were excluded from further analysis. Five lesions had less than 5mm of visible tumor on biopsy and were also not considered for molecular analysis. One of these lesions (lesion L11) had sufficient tumor (4.5 mm) for limited histologic analysis. A total of 48 lesions were subjected to combined histologic and molecular analysis. Generally, more than one biopsy was selected for a lesion if it contained large volumes of tumor conducive to laser capture microdissection or if multiple biopsies from that lesion displayed varying histologic phenotypes or adverse pathologies, such as Gleason score or perineural invasion. Using maps created from the RP surgical samples, we verified that every major region of residual tumor was in the same anatomical region as a biopsy acquired prior to treatment.

### Histology

5-micron serial sections of formalin-fixed paraffin-embedded (FFPE) biopsy tissues were stained with H&E, stained with antibodies by immunohistochemistry, or cut onto polyethylene naphthalate membrane slides (MicroDissect GmbH) for laser capture microdissection (LCM). For LCM, additional glass slides were cut after every five membrane slides for additional H&E and IHC stains to serve as references.

For IHC, slides were stained using validated protocols with antibodies against androgen receptor (AR), prostate specific antigen (PSA), glucocorticoid receptor (GR), ERG, PTEN, synaptophysin (SYP), PIN-4, and Ki-67 on an IP FLX autostainer (Biocare Medical). Glass slides containing tissue sections were baked for 30 minutes at 60°C. Following deparaffinization in xylenes and rehydration through graded alcohols, antigen retrieval was performed using a NxGen Decloaker (Biocare Medical) at 110°C for 15 minutes in Tris-EDTA Buffer (Abcam; ab93684), pH 9.0 for PSA, PTEN, and Synaptophysin or Diva Decloaker (BioCare Medical; DV2004MX) for AR, GR, ERG, PIN-4, and Ki-67. Next, a thin border was drawn around the edges of each glass slide using a PAP pen. 300 μL of primary antibody solutions were prepared and incubated at room temperature as follows: anti-AR clone D6F11 (Cell Signaling; 5153) diluted 1:200 into Renoir Red diluent (Biocare; PD904) for 30 m, anti-PSA (DAKO; IR514) ready-to-use (RTU) for 30 m, anti-SYP (Dako; M7315) diluted 1:200 into Renoir Red for 30 m, PIN-4 cocktail (CK5/14 + p63 + P504S) (BioCare Medical; PPM225DSH) RTU for 1 h, anti-PTEN clone D4.3 (Cell Signaling; 9188) diluted 1:100 into Renoir Red for 1 h, anti-Ki-67 clone D2H10 (Cell Signaling; 9027) diluted 1:500 into Cell SignalStain diluent (Cell Signaling; 8112) for 30 m, anti-GR clone D6H2L (Cell Signaling; 12041) diluted 1:400 into Renoir Red for 30 m, or anti-ERG clone EPR3864 (Abcam; ab92513) diluted 1:200 into SignalStain diluent for 30 m. Secondary detection was achieved with Mach 2 (for PIN-4) (Biocare Medical; MRCT525) or Mach 4 (Biocare Medical; M4U534H) polymer and/or probe (all other antibodies) for 30 minutes. Chromogen development was achieved with either Vulcan Red Fast Chromogen (Biocare Medical; FR805) or Betazoid DAB (Biocare Medical; BDB2004) and counterstained with CAT hematoxylin (Biocare Medical; CATHE) diluted 1:2 into distilled water. PIN-4 stained slides were air dried; all other slides were dehydrated through graded alcohols into xylene. Slides were mounted using Permount (Thermo Fisher).

For each stain, internal or external controls were employed to validate staining. For anti-AR, nuclear staining of luminal cells in normal prostatic tissue were positive controls, while ADT-treated tissue was used as a negative control. For anti-PSA, normal prostatic tissue was the positive control and non-prostate tissue was negative control. For anti-PTEN, normal tissue was used as the positive control and tissue previously confirmed to be genomically deleted for *PTEN* was the negative control. For anti-GR, endogenous staining in normal prostatic tissue was the positive control and FFPE-embedded LREX GR-knockout cells (kind gift of Charles Sawyers and David Wise) were negative controls for GR. For anti-SYP, pancreatic tissue (sourced from the NCI Laboratory of Pathology) was used as the positive control and PSA-positive prostate adenocarcinoma was the negative control. For anti-Ki-67, lymph node tissue invaded by MYC-amplified prostate cancer was used as the positive control and normal prostate luminal cells were the negative controls. For anti-ERG, normal prostate luminal cells were the negative control, and endogenous staining by endothelial cells was used as the positive control.

Calls of PTEN reduction on the focus or per level were made on the basis of least 5% of cancer cells showing reduced levels of PTEN intensity relative to normal glands or stromal cells in the same tissue. Calls of nuclear ERG expression on the focus or patient level were made on the basis of any cancer cell displaying positive nuclear ERG staining.

After mounted slides were fully dried, residual mounting media was removed using xylene. Slides were then digitized on Carl Zeiss AxioScan.Z1 microscope slide scanner equipped with a Plan-Apochromat 20× NA 0.8 objective, 266% LED intensity, 200 μs exposure time. Tissue images were acquired using ZEN Blue 2012 (Zeiss) with objective/magnification and pixel:distance calibrations recorded within the scanned CZI file. Digitized slides are pending deposit.

### Laser capture microdissection

Areas of interest on one or more biopsy tissues were identified by two genitourinary pathologists (R.T.L. and H.Y.) based on concordance or discordance with diagnostic pathology reports from each patient, the size of the tumor lesion estimated by mpMRI, adverse pathologic features identified on H&E tissues, or differential staining identified by IHC. Up to four areas per biopsy slide were annotated digitally on scanned H&E or IHC slides using ZEN Browser (Zeiss) and displayed on a second monitor for reference during LCM.

PEN-membrane slides were briefly baked, deparaffinized, rehydrated and stained with Paradise stain (Thermo Fisher) according to the manufacturer’s protocol. Approximately 10,000-50,000 tumor cells per ROI were captured from serial sections using an ArcturusXT Ti microscope onto CapSure Macro LCM Caps (Thermo Fisher) using the infrared capture and ultraviolet cutting lasers. Adjacent stromal tissue that incidentally captured was ablated using the UV laser. Micrographs of each cap were taken after each LCM session and cross-referenced against reference slides to verify the regions captured. Normal DNA was acquired from benign areas not involved with cancer, either from biopsies previously determined to be tumor-free, or from the post-treatment (surgical) specimen. After LCM, each captured tumor area was re-reviewed by two blinded genitourinary pathologists (H.A.S. and R.T.L.) to determine the highest Gleason pattern in the captured region, the percentage of the captured region corresponding to Gleason patterns 4 and 5, and whether the captured area contained ductal morphology, cribriform architecture, intraductal carcinoma of the prostate, diminished PTEN staining, or positive nuclear ERG staining.

### DNA extraction

Whole blood collected in K_2_EDTA tubes was centrifuged at 22°C for 20 minutes at 300 × *g*. After removal of the plasma layer, the buffy coat interface was transferred to a new tube and frozen at −80°C. gDNA was extracted from 10-100 μL of buffy coat using the DNeasy Blood and Tissue Kit (Qiagen) following the manufacturer’s protocol with two elutions of buffer AE (100 μL per elution). gDNA from 500 μL of preserved saliva was extracted using the prepIT L2P Kit (DNA Genotek) following the manufacturer’s instructions.

DNA was extracted from LCM tissues by using a clean scalpel to excise the LCM cap polymer and immerse it in Buffer ATL from the QIAamp DNA FFPE Tissue Kit (Qiagen). The initial proteinase K digestion step was performed overnight, and the remainder of the extraction protocol was followed according to the manufacturer’s instructions. DNA from blood, saliva and tissues were quantified using Picogreen reagent (Thermo Fisher).

### Sequencing and data processing

20-200 ng of double-stranded genomic DNA from tumor or normal regions was sheared (Covaris) and prepared into libraries using the SureSelect Human All Exon V7 Low Input Exome kit (Agilent) for whole-exome sequencing (WES). Pooled libraries were sequenced with 150 cycles paired-end plus 8 indexing cycles on either a HiSeq 4000 (Illumina) or a NovaSeq 6000 (Illumina). 200 ng of double-stranded genomic DNA from blood or saliva was sheared (Covaris) and prepared into dual-barcoded libraries using the Illumina TruSeq Nano kit and sequenced with 150 cycles paired-end plus 16 dual-barcoding indexing cycles on a NovaSeq 6000 for whole-genome sequencing (WGS). Raw sequencing data has been deposited in the Database of Genotypes and Phenotypes (dbGaP) accession number phs001938.v2.p1.

FASTQ files from WES were filtered to remove read pairs flagged as failed by sequencer. Pass-filter reads were then trimmed using SureCall Trimmer version 4.0.1 (Agilent) and aligned at the lane level with the Burrows Wheeler Aligner BWA-MEM version 0.7.17^21^ to version hg19 of the human genome (b37 with decoy chromosomes). The SAM alignment files were coordinate-sorted and duplicate-marked using PICARD version 2.18.27 SortSam and MarkDuplicates then quality score recalibrated using version 4.1.3.0 of the Genome Analysis Toolkit (GATK) BaseRecalibrator and ApplyBQSR. Lane-level BAM files were merged using PICARD MergeSamFiles and duplicate-marked at the sample level again using PICARD. On-bait capture efficiencies and library complexity were determined using PICARD CalculateHsMetrics.

FASTQ files from WGS were filtered to remove read pairs flagged as failed by the sequencer. Pass-filter reads were then trimmed using Trimmomatic version 0.39 and aligned at the lane level with the Burrows Wheeler Aligner BWA-MEM version 0.7.17 to version hg19 of the human genome (b37 with decoy chromosomes). All post alignment steps were performed using the Spark hyperthreading implementation of GATK 4.1.3.0. The SAM alignment files were coordinate-sorted using GATK MarkDuplicatesSpark and tag-set with PICARD SetNmMdAndUqTags. Base quality score recalibration was performed using GATK BQSRPipelineSpark, and then all lane-level BAM files were merged using PICARD MergeSamFiles. GATK MarkDuplicatesSpark and PICARD SetNmMdAndUqTags was run a second time on the merged, sample-level BAM file. WGS coverage metrics and library complexity were determined using PICARD CalculateWgsMetrics.

### Somatic variant calling

Somatic point mutations and indels were called on intervals from the Agilent bait design BED files using MuTect2 (part of the GATK4 package), first by running MuTect2 in tumor-only mode on all of the normal tissue BAM files individually, with the parameters disable-read-filter set to MateOnSameContigOrNoMappedMateReadFilter and max-mnp-distance set to 0. The “padded” bait BED file was used as the interval file of covered regions for all MuTect2 and filtering steps. Each of the output VCF files from this analysis of normal BAMs was merged into a database using GATK GenomicsDBImport with the setting merge-input-intervals set to true, and then generating a panel of normal from the database using GATK CreateSomaticPanelOfNormals. The WGS BAM files from buffy coat or saliva gDNA was subsetted to serve as a second matched normal using PICARD FilterSamReads with the Agilent bait design BED intervals file. MuTect2 was run in somatic mode on each tumor BAM paired with the normal WGS-subsetted BAM from the patient and the somatic panel of normals with af-of-alleles-not-in-resource set to 0.0000025 to exclude sites present in gnomAD and disable-read-filter set to MateOnSameContigOrNoMappedMateReadFilter. GetPileupSummaries and CalculateContamination were used on each tumor BAM file and the resultant contamination table was used to filter somatic mutations using FilterMutectCalls. CollectSequencingArtifactMetrics and FilterByOrientationBias were used to further filter mutations for 8-oxoG artifacts using the settings -AM G/T -AM C/T. These pass-filter mutations were then functionally annotated using Oncotator version 1.9.70 (database version April052016), while a second set of mutations that failed any of the filters were functionally annotated into a separate file. The number of small nucleotide variants (SNVs) reported per tumor focus is the number of pass-filter, coding, non-synonymous mutations (*i.e*. missense, nonsense, frameshift, splice-site, in-frame deletion/insertion, start/stop-codon, and *de novo* start mutations) that are further filtered to allow for no more than 10% strand bias at each site.

With limited power of this cohort to discover new mutations, variants considered in unbiased univariate analyses were curated first by only selecting the pass-filter mutations that overlapped with those deposited in COSMIC, and then further limiting those genes that also overlapped with a series of 727 cancer-related genes frequently mutated in prostate cancer and other human tumors ^22,23^. Mutations reported in large prostate cancer cohort studies^24,25^ were backfilled if not yet in COSMIC. Mutations considered in curated and pathway analyses were further classified as gain-of-function, loss-of-function, or hotspot mutations by manual interrogation in the Precision Oncology Knowledge Base (OncoKB, http://www.oncokb.org).

Underlying processes of mutational signatures were estimated from WES data using the deconstructSigs^26^ module for R version 3.5, using the Nature 2013^27^ signatures as the reference. All pass-filter synonymous and non-synonymous mutations were considered for this analysis, provided that they had less than 10% strand bias, were covered by the tumor and normal sample by at least 16× coverage, and were greater than 2% variant allele fraction in the tumor.

Somatic copy number alterations (SCNAs) were called across genomic intervals specified by the Agilent library design BED file with variable resolution depending on bait spacing. The design BED file was preprocessed with GATK PreprocessIntervals, with bin-length set to 0 and interval-merging-rule set to OVERLAPPING_ONLY. The BED file was also annotated with GC content using AnnotateIntervals. These interval files were used in all copy number calling steps. Read counts were first obtained from all tumor and normal BAM files using GATK CollectReadCounts. The normal read count files were compiled into a panel of normal using GATK CreateReadCountPanelOfNormals, excluding cases that had read depth coverage outside the 95% confidence interval for the dataset. The panel of normals was used to smooth read counts across all samples using GATK DenoiseReadCounts. WGS matched-normal BAM files subsetted for the region of coverage of the WES bait library were then processed with GATK CollectAllelicCounts to identify regions of potential LOH. GATK CollectAllelicCounts was also applied to each tumor BAM file. GATK ModelSegments used smoothed read counts from each tumor BAM along with the paired normal/tumor allelic counts for generating copy number estimates that were then called using GATK CallCopyRatioSegments. Finally, potential noise from each WES-derived SCNAs SEG file was removed setting small segments to zero if the size of the segment divided by the number of segments was less than an *a priori* determined cutoff value of 750. All zero-value segments were merged using bedtools version 2.29.0.

Determination of recurrent regions of genomic gains or losses, as well as discrete gene-level somatic copy number changes, were performed using GISTIC 2.0.23 on the GenePattern platform (module version 6.15.28). The cleaned focus-level SEG file was used as input with the following parameters: focal length cutoff set to 0.50, gene GISTIC set to yes, confidence level set to 0.90, cap value set to infinite, broad analysis set to on, max sample segments set to 10,000, arm peel set to no, and gene collapse method set to extreme. Post-GISTIC log_2_-copy number ratio threshold values were used for calling discrete gene-level calls as follows: > 1.3 = 2-copy gain; 0.1 to 1.3 = 1-copy gain; −1.3 to −0.1 = 1-copy loss; < −1.3 = 2-copy loss. Chromosome arm-level changes followed the same threshold scheme per tumor focus, provided that at least 50% of the chromosome arm was affected by the change. Focal regions of significant gains or losses (peaks) were considered both by the magnitude of the log_2_-copy number change and the *P* value of the 2-sided Fisher’s exact test when comparing ER to INR cases. Percent genome altered (PGA) was calculated for each focus by filtering the whole-genome estimated copy number SEG file for only those log_2_-copy number ratios greater or less than ±1, adding the size of each segment, and dividing the sum by 3,095,677,412, the size in nucleotides of the reference genome. Graphical depictions of mutations for each focus/lesion/case (Oncoprints) were generated using Oncoprinter ^28^.

For consolidating genomic data from individual foci to lesions, a pre-determined threshold was set for a minimum number of foci to harbor that alteration to call a particular copy number event for the entire lesion. For lesions sampled by 7 or 8 foci, the threshold was 3; for 3, 4, 5 or 6 foci the threshold was 2, and for 1 or 2 foci the threshold was 1. For each focus, per-gene somatic copy number calls from GISTIC (*i.e*. −1, −2, 0, 1 or 2) were averaged across all foci from each lesion. If the absolute value of that average was greater than the pre-determined threshold, the averaged value was rounded to the nearest integer. For point mutations, the lesion was considered to harbor the mutation if any number of foci harbored it. This approach allowed us to conservatively call copy number changes and mutations with similar sensitivities as bulk sequencing of non-microdissected tumor tissue. Univariate affected gene analyses considered any past-threshold somatic copy number alteration or mutation once consolidated at the lesion level. Similarly, genes with biallelic events were re-aggregated at the lesion level based on the same foci consolidation scheme. The log_2_-copy number changes of significantly altered focal peaks (gains or losses) were averaged across foci per lesion and univariate analyses performed on the aggregated data. For SNVs and PGA, values were averaged across all foci consolidated into a lesion. For mutational signatures, the proportion of each signature was averaged across foci in the same lesion. After all aggregation, lesion level was used directly for patient-level data except in cases where there were multiple lesions per patient; the index lesion served as the representative lesion for those patients.

For determining affected pathways at the patient or lesion level, the patient or lesion consensus effect for each gene (either mutation or copy number change) was considered individually as a gain-of-function and/or loss-of-function event based on the effect predicted in OncoKB. Genes were organized into a subset of cancer-related pathways^29,30^, and a pathway was reported affected in a lesion or case based on whether any genes in each group displayed the predicted effect to perturb that pathway. For the cell cycle, epigenetic, MYC, NOTCH and WNT pathways, only 1 gene alteration counted towards the pathway being altered. For the DNA repair, PI-3K, and RAS-MAPK pathways, two genes had to be affected.

### Germline variant calling

Germline variants were identified from buffy coat or saliva WGS BAM files individually by running GATK HaplotypeCaller with ERC set to GVCF and referencing dbsnp version 138. All VCF files were joined into a cohort by running GATK CombineGVCFs and genotyped jointly with GATK GenotypeGVCFs. All discovered variants were subjected to sequential recalibration. SNVs were first recalibrated using GATK VariantRecalibrator with the following parameters: -- resource:hapmap,known=false,training=true,truth=true,prior=15.0 hapmap_3.3.b37.vcf -- resource:omni,known=false,training=true,truth=false,prior=12.0 1000G_omni2.5.b37.vcf -- resource:1000G,known=false,training=true,truth=false,prior=10.0 1000G_phase1.snps.high_confidence.b37.vcf -- resource:dbsnp,known=true,training=false,truth=false,prior=2.0 dbsnp_138.b37.vcf --an QD -an MQ-an MQRankSum -an ReadPosRankSum -an FS -an SOR -an DP -mode SNP with outputs applied using GATK ApplyVSQR with the following parameters: -ts-filter-level 99.5 -mode SNP. Similarly, indels were then recalibrated using GATK VariantRecalibrator with the following parameters: -- resource:mills,known=false,training=true,truth=true,prior=12.0 Mills_and_1000G_gold_standard.indels.b37.vcf -- resource:dbsnp,known=true,training=false,truth=false,prior=2.0 dbsnp_138.b37.vcf -an QD -an DP - an FS -an SOR -an ReadPosRankSum -an MQRankSum --max-gaussians 4 -mode INDEL with outputs applied using GATK ApplyVSQR with the following parameters: -ts-filter-level 99.0 –mode INDEL. The VCF file of recalibrated SNPs was annotated in the “id” column with the rsid of SNPs from dbSNP using bcftools (Samtools version 1.3.1).

Germline mutations were then annotated using Oncotator version 1.9.70 (database version April052016), considering only obvious loss-of-function mutations (*i.e*. frameshift, nonsense) identified by filtering for mutations overlapping with the cancer gene census germline mutation database. For a mutation to be considered as a germline mutation it had to be considered heterozygous by HaplotypeCaller and be covered at greater than 15× coverage.

### Tumor phylogeny estimation

To estimate the evolutionary path of each tumor and identify tumor subpopulations, we aggregated gene-level somatic mutation and somatic copy number data by patient. For somatic mutations, we compiled a list of all high-quality (no strand bias), high-depth (at least 100× coverage), pass-filter coding mutations for each patient. In cases where the number of independent mutations across all foci was less than 50, stringency restrictions were lessened, starting with including noncoding mutations, then relaxing strand bias, then by decreasing the depth requirements to half of the average depth of each sample but no less than 16× coverage. The rationale for this conservative selection process was to start with the mutations that would be the most informative for inferring clonality from variant allele frequency. Then for each independent mutation within each patient, the same chromosomal coordinate was assessed against the raw list of variants and backfilled for the remainder of the tumor foci. This Bayesian process allowed previously filtered but shared mutations to be used as evidence for supporting common ancestry in conjunction with high quality mutations in other samples from the same patient, while also allowing unshared high quality mutations as evidence for subclonal populations.

For somatic copy number alterations, per-sample determination of gain or loss segments by GATK resulted in close (but not exact) boundaries for the same event across multiple foci from the same sample. To prevent this noise from supporting spurious subclones during analysis, the cleaned SEG file processed with GISTIC 2.0 identified several hundred broad or focal peaks with harmonized boundaries for each sample. Copy number gains and losses were then re-called based on the unified segment boundaries within each case, allowing for both shared and unshared SCNA events across foci. A histogram of variant allele frequencies for each sample was used to estimate the clonal and subclonal mutations for determining tumor purity and inferring major alleles, minor alleles, and cellular prevalence of SCNA segment.

Prior to running PhyloWGS^31^ version 1.0, the create_phylowgs_inputs tool was used to parse informative SCNAs (CNVs) and mutations (SSMs) from multiple VCF and SEG files into a single set of inputs. The SSM file was then parsed to re-annotate each mutation’s coordinate with the functional annotation from Oncotator. Finally, the multievolve script was used to build phylogenetic trees. For each case, 40 MCMC chains were run simultaneously with 1,000 burn-in samples and 2,500 MCMC-samples, for a total of 100,000 possible trees considered for each patient. After tree generation, chains were merged using the write_results tool, and the mutation and tree JSON files were parsed to select the tree with the most negative log likelihood score. Although multiprimary trees were allowed in the output, only 1 out of 37 trees generated a polytumor tree, suggesting that 100,000 potential trees allowed for sufficient estimation of subclonal reconstruction without defaulting to multiple independent clones.

The best scoring tree was then pruned using a predefined set of rules^5^ to conservatively decrease the number of potential subclones. If any given node did not have at least 5 SNVs or SSMs assigned to it, it was merged with its sibling node with the greatest number of events. If that had no siblings, it was merged with its most immediate ancestral node, unless it was a direct descendant of the germ/normal node with no descendants, in which case it was eliminated. The only nodes with fewer than 5 SNVs/SSMs were the parent cancer node at least one descendant that could not be merged. The one tree that suggested a polytumor evolutionary path before pruning remained a polytumor after pruning, suggesting sufficient evidence supported those node assignments.

The mean value of the cellular prevalences of all alterations assigned to the node served as the cellular prevalence for each node in the tree. Graphical depictions of node sizes on tree figures are proportional to the cellular prevalence. Shannon diversity indices (SDI) were calculated using the conventional formula for the SDI (*H*), in which

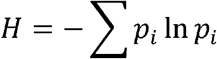

where *p*_*i*_ is the proportion of each node’s cellular prevalence in the entire population. The script for processing PhyloWGS data has been deposited at https://github.com/CBIIT/lgcp.

### Automated image analysis

Whole biopsy slide CZI files of slides stained with antibodies against AR, PSA, Ki-67, SYP, and GR were imported into Definiens Developer XD 64. The magnification for each analysis was set to 20×, with 0.21 µm/pixel for all solutions. IHC stain was identified as brown chromogen. The tumor cells were counted using their nuclear stain. Composer magnification was set to 6× with 12 training subsets, with training segmentation at level 9 for all stains except for SYP, which was segmented at level 8. Segments were classified as tumor or stroma, with normal glands excluded. Within the tumor pattern, cellular analysis magnification was set to 10× with 12 training subsets, and within nuclear detection, the hematoxylin threshold was set at 0.05-0.2 and the brown chromogen was set between 0.15 and 0.3, with typical nuclear size set to 30 µm. For PSA and SYP, nuclei were used to simulate cells, where cytoplasm was estimated at a maximum distance of 4-8 µm around the nucleus.

For Ki-67, SYP and GR, an additional filter was used to exclude nuclei between 15-25µm when non-specific tumor calling was present. Nuclear classification for AR was low vs. medium at 0.7 and medium vs. high at 0.95. Cellular classification for PSA was no expression vs. low at 0.2, low vs. medium at 0.4, and medium vs. high at 0.5. Nucleus classification for Ki-67 was low vs. medium at 0.65 and medium vs. high at 0.86. Cellular classification for SYP was no expression vs. low at 0.2, low vs. medium at 0.4 and medium vs. high at 0.5. Nuclear classification for GR was low vs. medium at 0.4 and medium vs. high at 0.6.

The total number of positively stained nuclei were reported, along with distribution of low-, medium-, and high-intensity stained nuclei for each tumor focus. A percent positive histology index (HI) score was calculated using a weighted average divided by the total number of nuclei, where index = [(1 × nuclei/cell stained low) + (2 × nuclei/cell stained medium) + (3 × nuclei/cell stained high)] ÷ (3 × total nuclei). The Shannon diversity index (SDI) was calculated as described above. The nuclei or cells quantified for each case were aggregated across blocks prior to calculating HI or SDI.

### The Cancer Genome Atlas (TCGA) analysis

Whole exome sequencing, clinical, pathological, and recurrence data from 188 tumors harboring Gleason patterns 4 and/or 5 (*i.e*. Gleason scores of 4+4=8, 4+5=9, 5+4=9 and 5+5=10) were accessed and downloaded from the National Cancer Institute Genomic Data Commons (http://gdc.cancer.gov) via authorized access project #9940 under dbGaP accession ID phs000178. Single sample whole-exome sequencing data was aligned and processed identically to the study cohort data for the purposes of identifying somatic mutations, germline mutations, somatic copy number alterations, SNV counts and PGA. ERG-fusion status and pre-operative PSA levels were included if available. Phylogeny estimation followed the same procedure as single-focus study samples, considering 100,000 possible tree structures for each tumor.

Whole slide images of frozen tumor sections used for TCGA molecular analysis were accessed on the NCI Genomic Data Commons and cross referenced against a prior publication^32^ for the presence of invasive cribriform carcinoma or intraductal carcinoma (IDC-P), as they were not distinguished. Cases previously annotated as ICC were re-reviewed by a genitourinary pathologist (R.T.L.) and further classified as IDC-P based on the presence of nuclear atypia, packed lumina, ducts with florid luminal carcinoma, and the number of glands involved with carcinoma, while excluding atrophic glands with flattened epithelia. For each call of IDC-P presence or absence, a confidence score was calculated based on these factors, the presence of adjacent invasive adenocarcinoma, and whether IDC-P was clearly visible on formalin sections from the same patient. Only cases with >99% confidence of IDC-P or no IDC-P were used in the validation cohort.

### Statistical analysis

Statistical analyses were performed with GraphPad Prism version 8, Microsoft Excel for Mac version 16, and SAS/STAT software version 14.1. Null hypothesis tests of associations (enrichments) between response and individual dichotomous factors were performed using a 2-sided Fisher’s exact test. Estimated odds ratios were calculated with 95% confidence intervals derived from inverted one-sided exact tests. Associations between factors were measured using Pearson or Spearman correlations. Comparisons of single factors between ER and INR cases were performed using Student’s or Welch’s *t* test or the Wilcoxon rank sum test. Comparisons of covariates within single factors between ER and INR cases were performed using the Cochran-Armitage test for trend. Logistic regression models including the four selected factors were constructed using forward and backward selection methods. Estimated probabilities were compared to the observed response proportions of the factor combinations. Sensitivity and specificity of dichotomized estimated probabilities were examined using receiver operating characteristic curves. Response probabilities were estimated as the same function of the factor values in an independent validation population. Genomic SDI scores were tested for Pearson correlations with these probabilities. The importance of each of the four factors in this correlation was assessed by fitting a new logistic regression model with the factor removed and comparing the probabilities to genomic SDI with the factor’s mean effect subtracted.

## RESULTS

### All cases are predominantly C class tumors with a paucity of coding point mutations in driver genes

In our clinical trial, 37 out of 39 patients completed the protocol of intense neoadjuvant ADT (inADT) of ADT plus enzalutamide for 6 months and underwent prostate surgery (Table 1). Prior to therapy, templated and MR/US-fusion targeted biopsies were obtained. We used laser capture microdissection to sample 141 histologically distinct tumor foci from these biopsies (median of 4 foci per patient) along with benign tissue for each case. We performed whole-exome sequencing on these microdissected foci to a mean on-target depth of 52.1× unique coverage and assessed the extent of whole-genome somatic copy number alterations and a spectrum of chromosomal number events and gene mutations frequently reported in prostate cancer (Supplementary Fig. 1). Germline alterations to cancer-related genes were detected by whole-genome sequencing of gDNA from saliva or buffy coats at a mean depth of 46.4× coverage (see Supplementary analysis). These 141 biopsies corresponded to 48 distinct MRI-contoured lesions (Supplementary Fig. 2a). Similar to previous studies assessing the genomic characteristics of intermediate and high-risk prostate cancer, the vast majority of mutation events’ potential driver genes were somatic copy number changes, consistent with prostate cancer’s classification as a C-class tumor ^33,34^. Interestingly, there was variable genomic heterogeneity between foci from the same MRI-defined lesion, as well as between lesions in the same patient (see Supplementary Figs. 1 and 2a). When these data were collapsed to single data points for each patient (Fig. 1a), the genomic landscape of recurrent somatic alterations was not significantly different than 188 high risk, Gleason pattern 4 and 5 tumors from the TCGA-PRAD^24^ dataset (Supplementary Fig. 3) when processed through the same bioinformatic processing pipeline (Fig. 1b, *P* = 0.098, Wilcoxon matched-pairs test). Individually, 27 of 28 curated genomic features did not show statistically significant differences (Chi-square test) between our clinical trial and the TCGA cohort, indicating that the high-risk population in our study is largely representative of locally advanced prostate cancers.

**Table 1.** Patient characteristics.

|  | *All patients* | *Exceptional responder* | *Incomplete or non-responder* |
| --- | --- | --- | --- |
| *N* | 37 | 15 | 22 |
| **Age (years), median (IQR)** | 62 (58–69) | 63 (58-71) | 62 (56-67) |
| **Race, *n* (%)** |  |  |  |
| **White** | 25 (68%) | 11 (73%) | 14 (64%) |
| **Black** | 6 (16%) | 3 (20%) | 3 (14%) |
| **Other** | 6 (16%) | 1 (7%) | 5 (23%) |
| **Baseline PSA (ng/ml), median (IQR)** | 9.31 (5.53–17.29) | 9.58 (5.90-21.37) | 8.05 (5.37-15.38) |
| **Baseline PSA density, median (IQR)** | 0.22 (0.15–0.37) | 0.22 (0.16-0.40) | 0.21 (0.15-0.36) |
| **Prostate vol. (ml), median (IQR)** | 40.0 (33.0-49.0) | 43.0 (35.0-56.5) | 36.5 (31.6-47.9) |
| **MRI tumor burden (cc), median (IQR)** | 3.44 (1.77-9.80) | 1.58 (1.27-3.07) | 6.99 (3.13-18.53) |
| **Pretreatment clinical stage, *n* (%)** |  |  |  |
| **T1** | 4 (11%) | 2 (13%) | 2 (9%) |
| **T2** | 6 (16%) | 5 (33%) | 1 (5%) |
| **T3a** | 15 (41%) | 5 (33%) | 10 (45%) |
| **T3b** | 7 (19%) | 1 (7%) | 6 (27%) |
| **T4** | 5 (14%) | 2 (13%) | 3 (14%) |
| **N1** | 11 (30%) | 3 (20%) | 8 (36%) |
| **ISUP Grade Group (biopsy), *n* (%)** |  |  |  |
| **2** | 4 (11%) | 0 (0%) | 4 (18%) |
| **3** | 5 (14%) | 2 (13%) | 3 (14%) |
| **4** | 11 (30%) | 7 (47%) | 4 (18%) |
| **5** | 17 (46%) | 6 (40%) | 11 (50%) |

**Figure 1.**
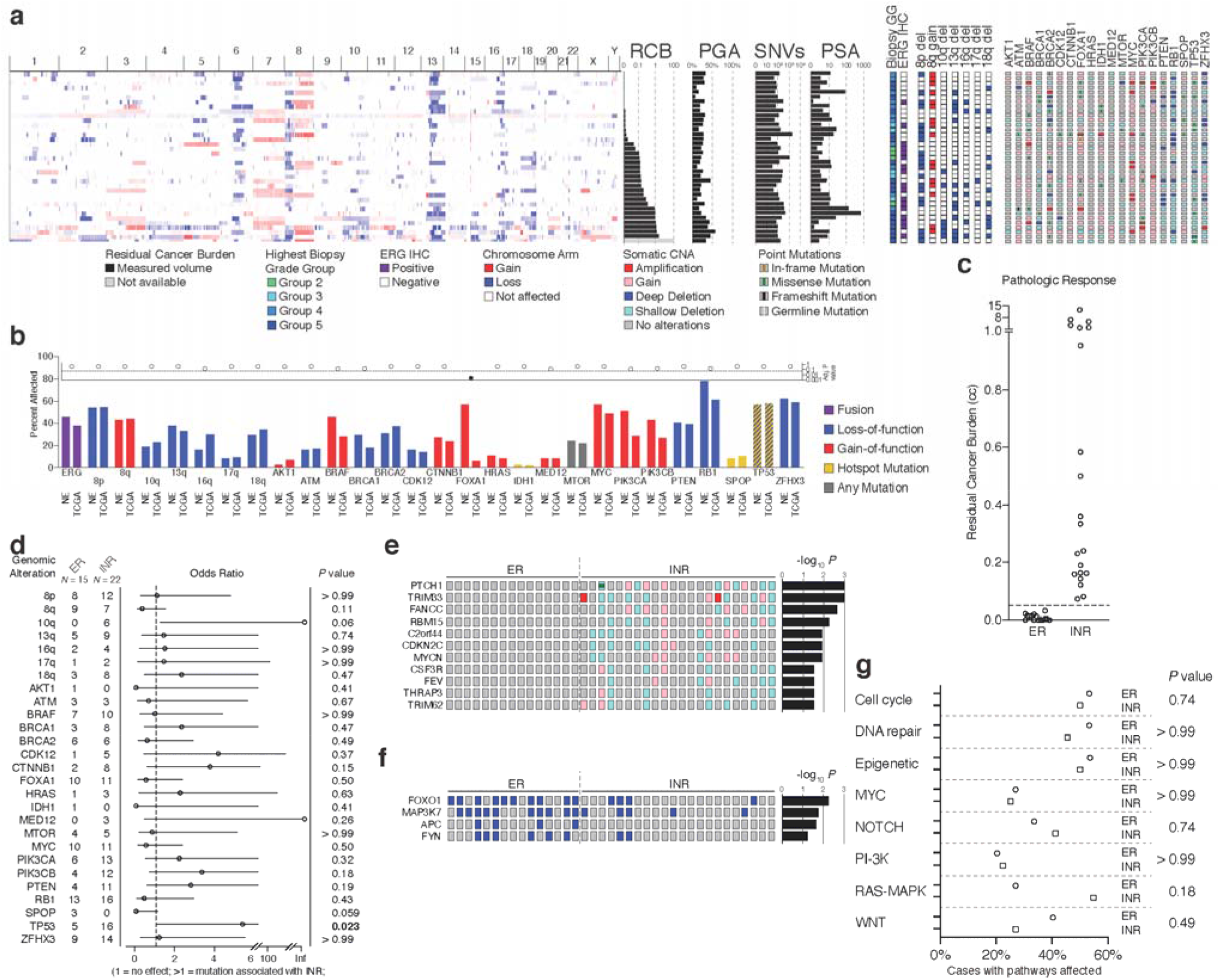
Genomic landscape of prostate tumors prior to treatment with neoadjuvant androgen deprivation therapy plus enzalutamide. (a) Genomic and clinical data are shown per patient (*N* = 37). One patient classified clinically as a non-responder underwent TURP instead of RP such that no calculation of RCB was possible. (b) Comparison of proportions for 28 genomic features across the current study cohort and the prostate cancer TCGA cases with Gleason scores 8-10 (*N* = 188). (c) Distribution of RCB values across 15 ER and 21 INR cases with a 0.05 cc cutpoint. (d) Frequency and odds ratio for 27 genomic features, comparing INR cases to ER cases. Odds ratio > 1 favors INR. (e) Oncoprint of genomic alterations observed exclusively in INR cases with *P* < 0.05 comparing INR to ER. (f) Oncoprint of biallelic events with *P* < 0.05 comparing INR to ER. (g) Grouping of altered genes into 8 pathways, with the proportion of affected cases with at least one or two genes altered per pathway. For (d-g), each factor was considered independently by a 2-sided Fisher’s exact test.

Following treatment we used the final radical prostatectomy specimen to assess local responses to therapy by calculating residual cancer burden (RCB), a validated intermediate endpoint for systemic therapy that has clinical significance for predicting recurrence after surgery ^14,15^. Based on prior neoadjuvant studies, the expected minimal residual disease (MRD) rate, and distribution of values, we selected 0.05 cc as the cut point (Fig. 1c) between exceptional responders (ER) and incomplete and nonresponders (INR) at the patient level, resulting in 15 ER patients and 22 INR patients. One patient whose tumor expanded during therapy underwent a TURP instead of radical prostatectomy and an RCB value could not be calculated. Within the INR group, registration of pre-treatment biopsies and imaging to final MRI and pathology identified 12 additional biopsy-confirmed lesions of which 10 exhibited exceptional pathological response, for a total of 25 ER and 23 INR MRI-visible lesions.

We then compared the frequency of 27 recurrent genomic events between the two groups by univariate analysis (Fig. 1d). Although arm-level loss of chromosome 10q was exclusive to the INR group and hotspot mutations to *SPOP* were only observed in the ER group, these did not reach statistical significance. As expected, somatic copy number losses and hotspot mutations to *TP53* were significantly enriched in the INR group (*P* = 0.023, Fisher’s exact test) at baseline (see Supplementary analysis). Using the imaging-responsive lesions from INR patients to sample the properties of sensitive tumors, similar trends were also observed at the lesion level (Supplementary Fig. 2b) with the finding that loss of chromosome 10q was significantly associated with poor response to inADT.

In a series of exploratory analyses, we also sought to identify perturbations to the genome that were associated with response to inADT. Using segmented copy number values from each focus of tumor in our entire cohort, we identified a series of 180 peaks using GISTIC and called gain or loss values within each peak (Supplementary Table 1), of which 5 were enriched in the INR group (*P* < 0.05, Fisher’s exact test). In addition to peaks identified by this analysis (Supplementary Fig. 4a), which included a portion of chromosome 21 interstitially deleted in a subset of *TMPRSS2-ERG* fusions (q22.2) and loss at chromosome 10q21 encompassing *PTEN*, analysis at the lesion level (Supplementary Table 2) identified a loss at chromosome 5q14.3-15 that was enriched in the ER group (Supplementary Fig. 4b).

Interrogating the combined effect of somatic copy number changes and mutations, we examined at the patient-(Supplementary Table 3) and lesion-level (Supplementary Table 4) whether any alteration to a series of 712 cancer-related genes was mutually exclusive to the ER or INR group. As shown in Figure 1e, these univariate analyses revealed that mutations and copy number alterations to the hedgehog pathway receptor *PTCH1* were only observed in INR cases (*P* = 0.0009, Fisher’s exact test) and were enriched in INR lesions (Supplementary Fig. 2c). Other genes exclusively altered in INR included the tumor suppressor *FANCC* and the oncogene *MYCN*. As the majority of these alterations included shallow gains or deletions, we repeated this analysis to identify any gene enriched with biallelic inactivations, either by two-copy deletion or mutation plus loss of heterozygosity. Four genes satisfied these criteria at the patient level, all enriched in ER cases (Fig. 1f), which included *MAP3K7* and the oncogene *FYN* (*P* = 0.016 and 0.042, respectively); both are located in a recurrently-deleted region of chromosome 6q15-21. *FOXO1* deep deletions were also enriched in ER, both at the patient and lesion level (Supplementary Fig. 2d). The lesion-level analysis also identified homozygous deletions or hemizygous loss plus point mutations of *NOTCH2* exclusively in the INR group (*P* = 0.046).

When we grouped driver oncogene amplifications, tumor suppressor deletions, and point mutations into curated cancer gene pathways^29^, a greater proportion of INR tumors accumulated mutations in the RAS/RAF/MAPK pathway, which was statistically significant at the lesion level (Fig. 1g and Supplementary Fig. 2e). Although DNA homologous recombination gene mutations (*BRCA1, BRCA2*, and *PALB2*) were observed in all cases, neither group was significantly enriched for DNA damage repair gene alterations, consistent with similar mutation type proportions and signatures between ER and INR patients (Supplementary Fig. 5a-d). Thus, both ER and INR tumors demonstrated features of high-risk tumors and significantly altered genomes but harbored relatively few differences between groups.

### Pathologic and histologic features distinguish exceptional from incompletely- and non-responding tumors

With a striking range of pathologic responses across the cohort, we explored whether clinicopathologic parameters can explain at least in part the differences we have observed. Although INR patient tumor burdens were greater at baseline (see Table 1), there was no difference between ER and INR patients either in their baseline serum PSA levels (Fig. 2a), or in the highest Gleason Grade Group from pre-treatment biopsies per patient (Fig. 2b) and per mpMRI-visible lesion (Supplementary Fig. 6a). On univariate analysis, the proportion of cases or lesions harboring cribriform architecture, ductal morphology, or Gleason patterns 4 or 5 did not distinguish ER from INR, but nuclear ERG and intraductal carcinoma (IDC-P) were significantly different (Fig. 2c-d and Supplementary Fig. 6b). In particular, both nuclear ERG and IDC-P were enriched in INR cases (*P* = 0.0020 and 0.013, respectively).

**Figure 2.**
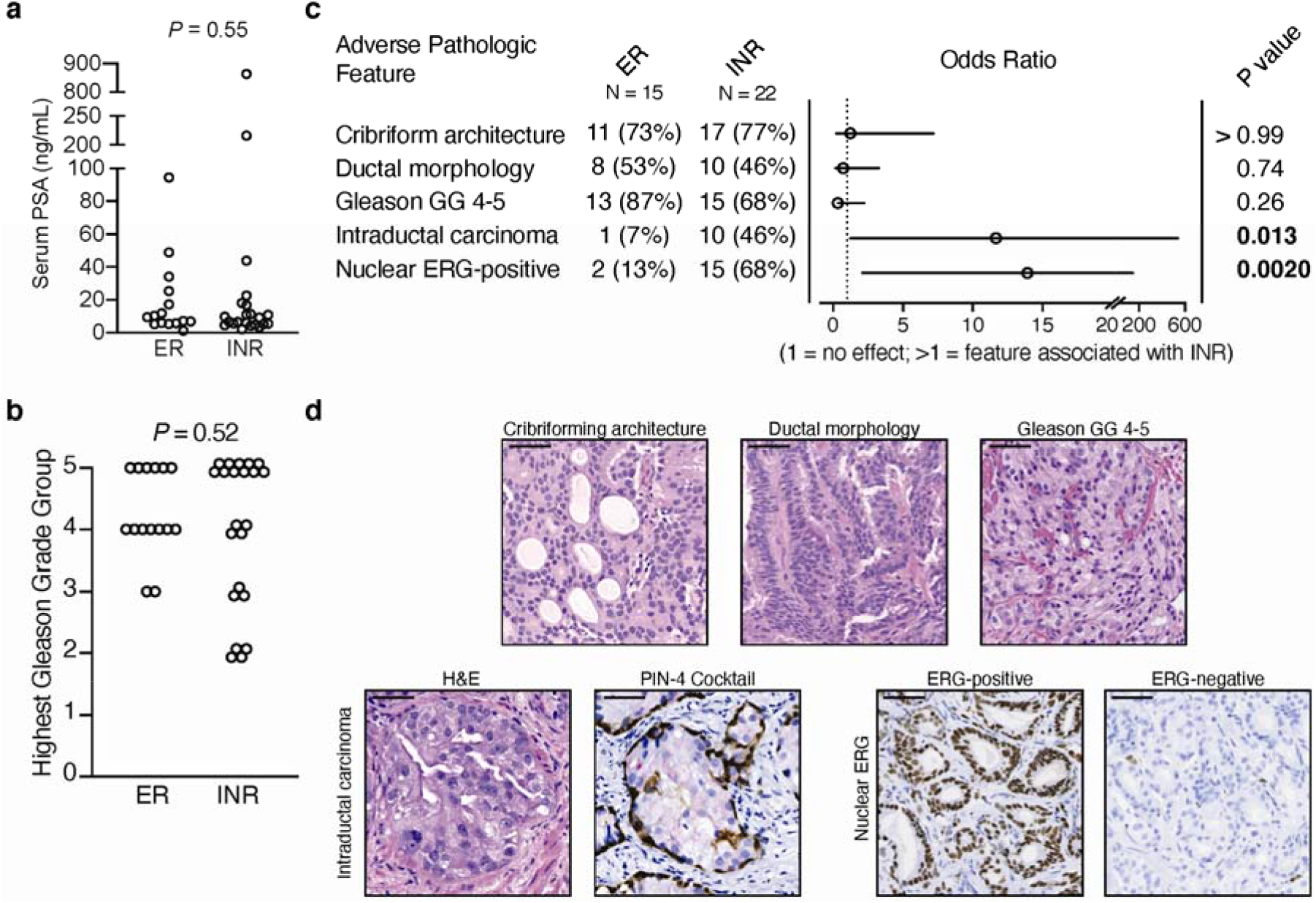
Comparison of clinicopathologic features between exceptional responder and incomplete and nonresponder cases. (a) Baseline serum PSA measurements comparing ER to INR. *P* = 0.55 by Mann-Whitney test. (b) The highest Gleason Grade Grouping (GG) of all pre-treatment biopsies for each case. *P* = 0.52 by the Cochran-Armitage test for trend for each GG between ER and INR. (c) Frequency and odds ratio for five histologic and pathologic features, comparing INR cases to ER cases. Odds ratio > 1 favors INR. (d) Representative micrographs of the adverse histologic features considered in (c). For intraductal carcinoma, PIN-4 cocktail stains basal cells with brown chromogen (antibodies against cytokeratin 5, cytokeratin 14, and p63) and luminal cells with red chromogen (antibody again alpha-methylacyl-CoA racemase). For nuclear ERG staining, endogenous ERG is observed in endothelial cells for ERG-negative cases. Scale bar: 50 μm.

As suggested by the frequency of *PTEN* alterations at the genomic level (see Fig. 1d), reductions in PTEN staining by immunohistochemistry were enriched (*P* = 0.0015, Fisher’s exact test) in INR vs. ER (Fig. 3a). We also observed intratumoral heterogeneity in the co-occurrence of PTEN-intact and reduced, or ERG-positive and negative, tumors. ER and INR cases (Fig. 3b) or lesions (Fig. 3c) were often uniformly positive or negative for both PTEN and ERG (scoring of 0), but the frequency of tumors with either heterogeneous PTEN or ERG staining (score of 1) or both PTEN and ERG being heterogeneous (score of 2) was greater in INR (Fig. 3d, *P* = 0.0059 and *P* = 0.0068, Cochran-Armitage test for trend, respectively).

**Figure 3.**
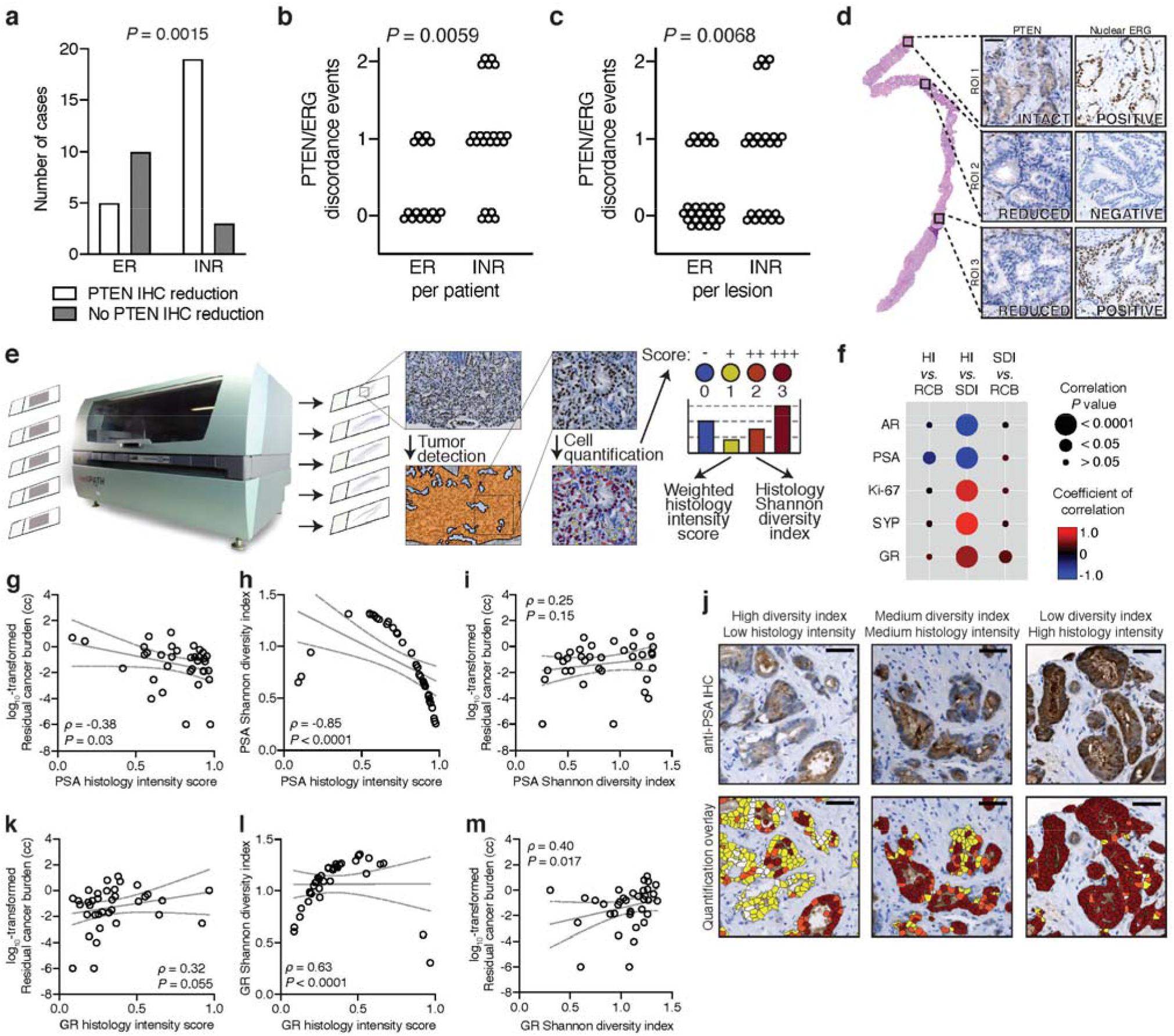
Observations of histologic diversity in baseline biopsies. (a) Enumeration of PTEN reduction observed by anti-PTEN IHC in ER and INR cases. (b-c) Per patient or per lesion enumeration when PTEN and/or ERG IHC is positive and negative within the same case. A score of 1 corresponds to either PTEN or ERG showing discordance. A score of 2 indicates both PTEN and ERG show heterogeneity within the same case. Statistical significance determined by the Cochran-Armitage test for trend. (d) A representative case demonstrated PTEN and ERG heterogeneity by IHC (score of 2), showing a region with intact PTEN and positive ERG staining (top), a region with reduced PTEN staining but negative staining for ERG (middle), and a region with reduced PTEN and positive ERG staining (bottom). All three insets are from the same biopsy sample. (e) Workflow for machine-based IHC quantification using an autostainer for reproducible staining conditions, followed by slide scanning and cell/nucleus quantification using the Definiens platform. (f) Bubble plot of IHC correlations between histology intensity (HI) score, residual cancer burden (RCB), and Shannon diversity index (SDI). The size of the bubble is inversely proportional to the Spearman rank correlation *P* value, and the color scale is red for more positive coefficients of correlation and blue for negative. (g, k) Correlation of the log_10_-transformed RCB for each case plotted against each case’s histology intensity score for anti-PSA (g) or anti-GR (k) IHC. (h, l) Shannon diversity index for anti-PSA (h) or anti-GR (l) IHC for each case plotted against each case’s histology intensity score for anti-PSA (h) or anti-GR (l) IHC. (i, m) Correlation of the log_10_-transformed RCB for each case plotted against each case’s Shannon diversity index for anti-PSA (i) or anti-GR (m) IHC. For (g, h, i, k, l and m), nonparametric Spearman correlation analyses *rho* (ρ) values are shown with their respective *P* values. Lines and bands represent linear regression lines and 95% confidence intervals, respectively. Due to the small number of points, the scatter plots in (h) and (l) appear parabolic with no known explanation, although the trend over most of the data suggests a linear relationship. (j) Representative regions of high, medium, and low staining diversity indices or low, medium, and high histology indices (respectively) of anti-PSA IHC. Scale bar: 50 μm.

Because the *TMPRSS2-ERG* fusion and reductions to PTEN tend to occur early during prostate tumorigenesis ^35^, our observation of their heterogeneity enriched amongst INR tumors at baseline raised the possibility that these tumors are more resistant to treatment in part due to a greater number of tumor subclones. As these subclones may display a wider array of prostate cancer phenotypes apparent *in situ*, we extended our histologic analyses to a panel of 5 additional immunostains on pre-treatment biopsies: androgen receptor (AR), prostate-specific antigen (PSA), glucocorticoid receptor (GR), synaptophysin (SYP), and Ki-67. Automated staining within the linear range of colorimetric development for each antibody, coupled with machine-learning based image analysis, resolved up to four subpopulations of tumor cells based on staining intensity, which in turn was used to derive weighted histology intensity (HI) scores and histology heterogeneity measures based on the Shannon diversity index (SDI, Fig. 3e). As depicted in Figure 3f, all stains demonstrated significant positive or negative associations between HI and SDI, indicating staining homogeneity at areas of high intensity (AR, PSA) or low intensity (Ki-67, SYP, GR), *i.e*. mostly all strong staining or mostly all weak staining. The histology scores for PSA were negatively correlated with the volume of residual disease (RCB) at the end of treatment (Fig. 3g). With the strong negative correlation between PSA HI and SDI (Fig. 3h), the positive correlation between PSA SDI and RCB (Fig. 3i) indicates that INR cases displayed more PSA heterogeneity as histologic intensity decreased (Fig. 3j). In contrast, ER cases were more uniformly strong for PSA staining, with a lower SDI at lower RCB. Unlike PSA, however, the histology index for GR was positively correlated with RCB (Fig. 3k), such that GR levels in tumor cells were generally and uniformly low but increased proportionally with RCB. GR intensity was proportional to its SDI (Fig. 3l), indicating that staining at greater intensities was associated with greater heterogeneity, which in turn was positively correlated with RCB (Fig. 3m). Consequently, GR intensity in tumor cells tended to be uniformly low except when displaying heterogeneity as its intensity increased, which was also proportional to RCB. As activation of GR is thought to be one of several potential bypass mechanisms for reconstituting AR activity under castrate levels of androgen ^36^, its increased staining heterogeneity at baseline may reflect a greater degree of tumor evolution in a subset of tumors leading to more diversity.

### Resistance to neoadjuvant enzalutamide is associated with greater genomic diversity at baseline

Given the increased histologic heterogeneity in the INR patient group, we next explored whether this phenomenon was also apparent at the genomic level. Superficially, we observed greater within-patient similarities when examining the whole-genome somatic copy number variation data of individual foci from exceptional responders than incomplete and nonresponders (see Supplementary Fig. 1. Using log_2_-copy number ratios derived by GISTIC across a defined set of 180 peaks (69 amplification, 111 deletion), Pearson correlation matrices confirmed that correlation coefficients were greater in patients with lower residual cancer burdens (Fig. 4a), irrespective of the number of sampled foci. Across all cases with at least two foci, there was a statistically significant negative correlation (ρ = −0.43; 95% C.I. −0.68 to −0.086; *P* = 0.014) between the median correlation coefficient of each case and the RCB volume following treatment (Fig. 4b) showing more intratumoral homogeneity in the exceptional responders. Although this observation could reflect more variable tumor purity in the INR group, another possibility is that the apparent heterogeneity reflects a more evolved tumor system with increasingly divergent subclones. As depicted in Fig. 4c, we therefore considered all callable, high-confidence mutations and boundary-harmonized somatic copy number events from every laser capture microdissected focus to identify tumor subpopulations via bioinformatic reconstruction of tumor phylogenies. We observed two general types of tree structures (Fig. 4d): 1) a relatively linear evolution path with few daughter subclones inferred from nodes; and 2) a more branched phylogenetic tree with multiple significant subclones. A depiction of each tree and supporting mutation/SCNA assignments is provided in Supplementary Figure 7.

**Figure 4.**
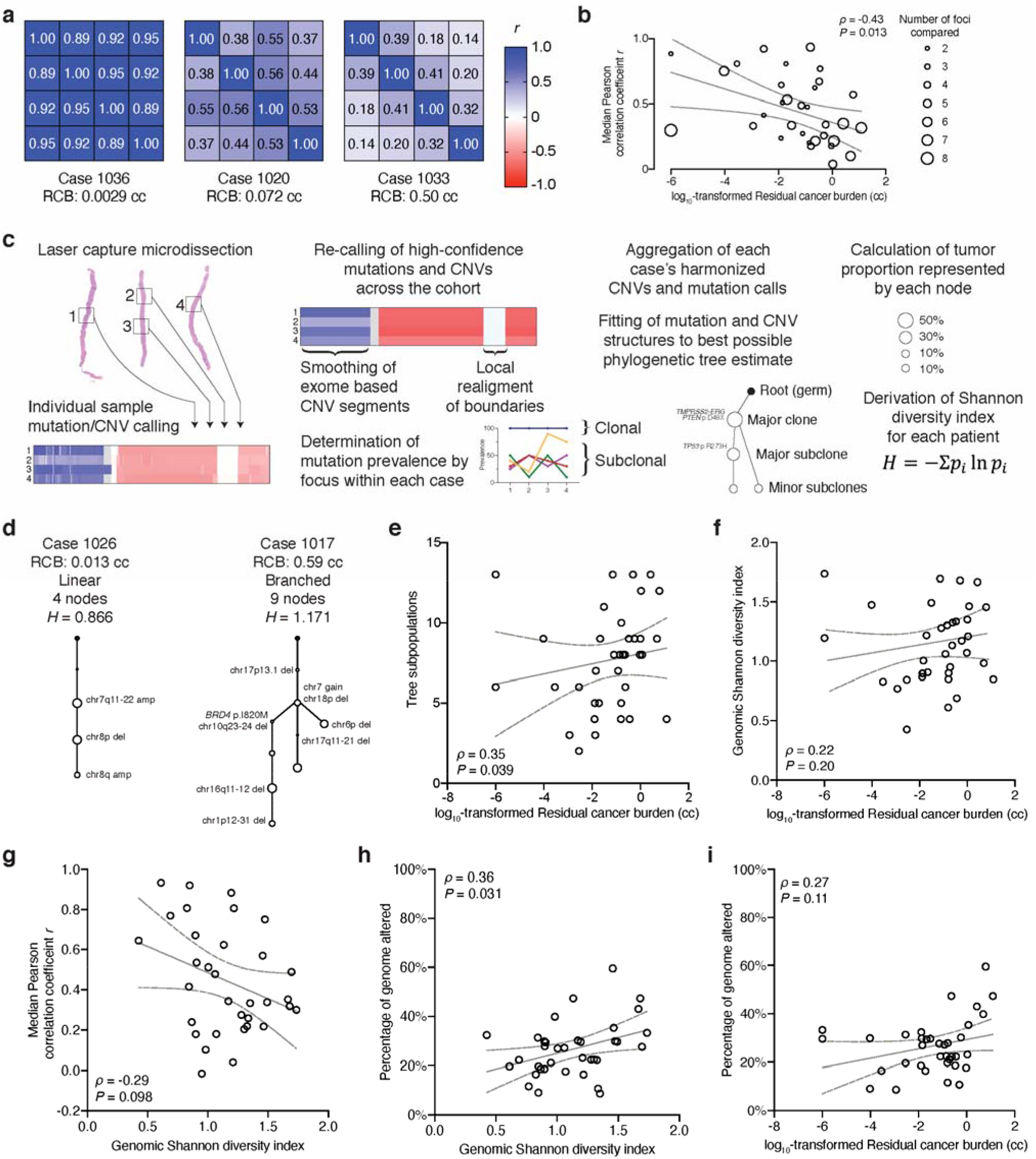
Measures of genomic diversity. (a) Correlation matrices from three representative cases with four tumors each showing the range of similarities amongst genomic gains and losses across 180 pre-defined regions. (b) The median Pearson correlation coefficient for each multi-region case is shown plotted as a function of the log_10_-transformed residual cancer burden (RCB) value. The number of foci dissected for each case is depicted by the size of the circle. (c) Schematic showing the workflow to harmonize multi-region sampling data and processing for phylogenetic tree reconstruction. (d) Representative linear and branched tree structures. (e) Scatter plot of the number of each phylogenetic tree’s nodes vs. the log_10_-transformed RCB for each case. (f) Scatter plot of the genomic Shannon diversity index (SDI) for each case vs. the corresponding log_10_-transformed RCB. (g) The median Pearson correlation coefficient for each multi-region sampled case from (b) plotted vs. the case’s genomic SDI. (h) The percentage of the genome altered (PGA) for each case plotted against each case’s genomic SDI. (i) PGA vs. the log_10_-transformed RCB for each case. For (b, e, f, g, h, and i), nonparametric Spearman correlation analyses *rho* (ρ) values are shown with their respective *P* values. Lines and bands represent linear regression lines and 95% confidence intervals, respectively.

Assessing these tree structures, we observed a greater proportion of branched trees in INR vs. ER cases (86% vs. 73%). The number of nodes in each tree (Fig. 4e) also showed a positive correlation with residual disease volumes (ρ = 0.35; 95% C.I. 0.0095 to 0.62; *P* = 0.039). When we used the prevalence of each tumor subpopulation to calculate the Shannon diversity index (SDI) for each tumor, we observed a modest positive correlation (ρ = 0.22; 95% C.I. −0.13 to 0.52; *P* = 0.20) between baseline SDI and post treatment RCB (Fig. 4f). Amongst cases with multiple foci, a modest inverse correlation was observed between copy number gain or loss inter-focal homogeneity and genomic SDI (ρ = −0.29; 95% C.I. −0.59 to 0.067; *P* = 0.098) indicating that these observations of intratumoral heterogeneity may be related to processes of evolutionary diversity (Fig. 4g). Moreover, as depicted in Fig. 4h-i, the percentage genome altered (PGA), a metric associated with tumor aggressiveness, was similarly positively correlated with both genomic SDI (ρ = 0.36; 95% C.I. 0.031 to 0.62; *P* = 0.03) and RCB (ρ = 0.27; 95% C.I. −0.071 to 0.56; *P* = 0.11). We therefore conclude from these observations that tumors with resistance to inADT may be associated with a greater number of major subclones that existed prior to initiation of therapy, and that sensitivity to inADT, even in tumors with driver alterations, may be at least partially explained by the decreased fitness conferred by tumors’ simpler evolutionary history.

To validate these findings, we first identified four independent factors from our univariate analyses that did not show perfect associations with each other (Fig. 5a) for constructing a multivariate model using logistic regression (Supplementary Table 5). The combination of these factors [presence of the *TMPRSS2:ERG*-fusion/nuclear ERG expression, deletion of at least half of chromosome 10q, loss-of-function alterations or hotspot mutations to *TP53*, and the presence of intraductal carcinoma (IDC-P)], allow up to 16 possible combinations, of which 9 were present in our dataset. As shown in Figure 5b, each combination is associated with a probability of being an INR case. This model (Fig. 5c) has an AUC of 0.8909 (*P* < 0.0001), and a cutpoint of 60% offered the best discrimination within the test cohort, classifying 30 of 37 cases correctly (*P* = 0.0002, Fisher’s exact test). This cutpoint also recapitulated the trend towards greater diversity seen initially, both in the number of nodes per tree and greater genomic SDI for each case with increased INR probability predicted by the model (Fig. 5d-f), demonstrating its potential utility in our trial cohort of diverse tumor profiles.

**Figure 5.**
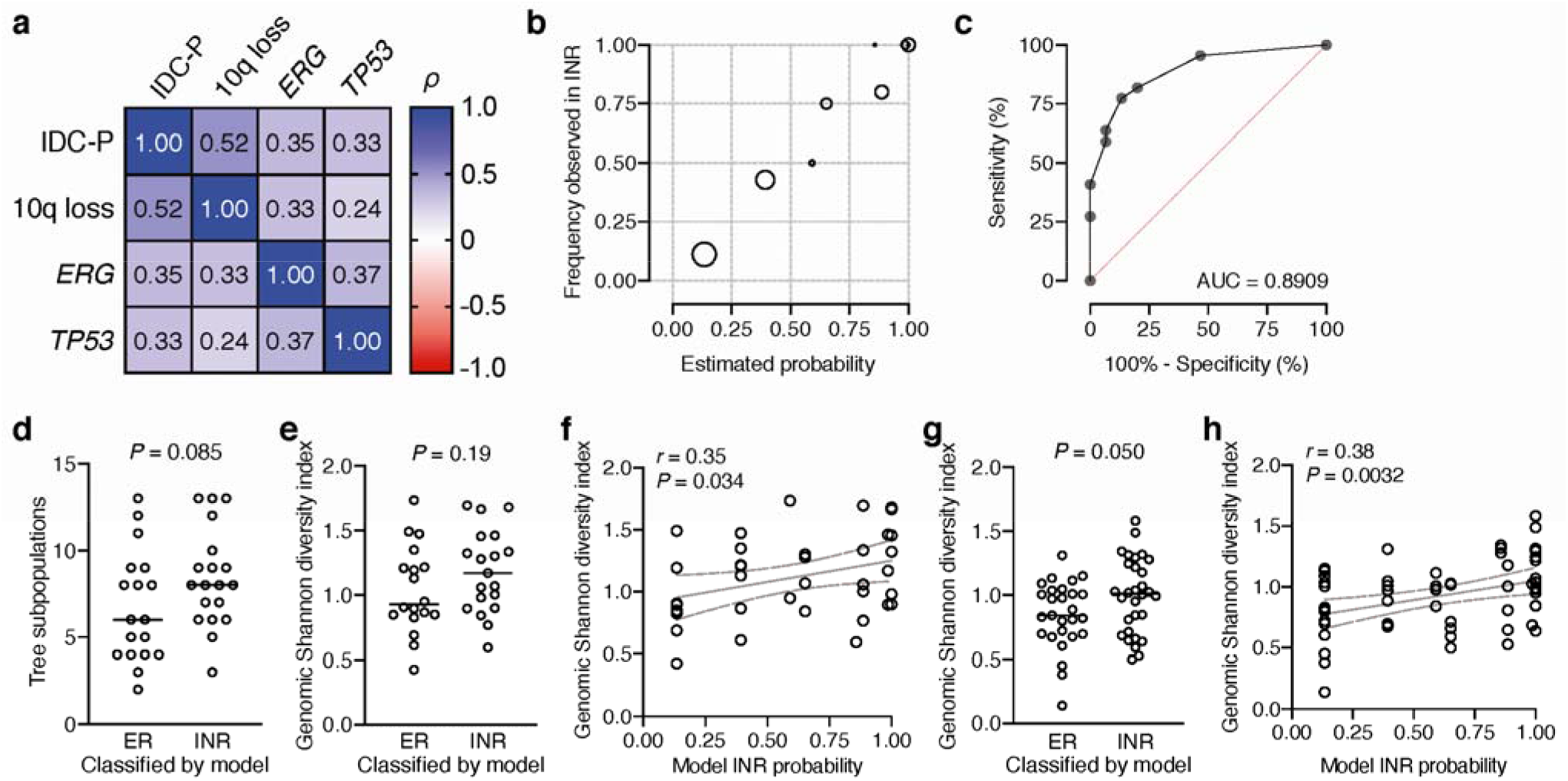
A multivariate model of poor response to intense neoadjuvant ADT is associated with genomic diversity. (a) Spearman correlation matrix of each binary factor (1 = presence, 0 = absence) across each of the 37 patients in the study cohort. IDC-P: presence of intraductal carcinoma; 10q loss: at least of half of chromosome arm 10q deleted hemizygously as determined by GISTIC; ERG: overexpression of nuclear ERG determined by immunohistochemistry; *TP53*: loss of function or hotspot mutations to *TP53*, including copy number loss as determined by GISTIC. (b) Bubble plot of the frequency of each 4-factor combination observed in the trial cohort and the proportion observed in the INR group. (c) Receiver operator characteristic (ROC) curve for the INR probabilities in distinguishing ER from INR in the trial cohort. (d-e) Re-application of the 4-factor model to trial patients and the number of tumor subpopulations (d) or the genomic Shannon diversity index (e) of each group after reclassification. Statistical significance determined using Mann-Whitney test for the number of tumor populations and Welch’s *t* test for genomic SDI. Line represents mean value. (f) Pearson correlation of the genomic SDI and INR probabilities for the trial cohort. (g) Genomic SDI of the TCGA validation cohort as classified as ER or INR by the 4-factor model. Statistical significance determined using Welch’s *t* test. Line represents mean value. (h) Pearson correlation of the genomic SDI and INR probabilities for the validation cohort.

Our validation group consisted of 188 Gleason score 8-10 cases from the prostate cancer TCGA cohort, of which 58 of the cases had complete data, which included the ERG-fusion annotation, 10q/*TP53* status and genomic SDI called via our bioinformatic pipeline, and presence or absence of IDC. The 60% INR probability cutpoint resulted in 27 cases classified ER and 30 classified INR. Consistent with our observations in the test cohort, TCGA cases classified as INR harbored a significantly higher genomic SDI than ER (*P* = 0.050, Welch’s *t* test, Fig. 5g), and the probability of INR across the validation group positively correlated (*r* = 0.38; C.I. 0.14 to 0.59; *P* = 0.0032) with genomic SDI (Fig. 5h). Consequently, greater genomic diversity in untreated prostate tumors is associated with increased likelihood of treatment resistance.

## DISCUSSION

Patients diagnosed with intermediate-to-high risk localized prostate cancer have variable outcomes, and in this clinical trial we found that pathologic responses to intense neoadjuvant ADT did not trend with classical clinicopathologic parameters observed at baseline, such as Gleason score or clinical stage. In this study, we used WES and IHC to identify potential molecular differences between exceptional responders and incomplete or nonresponders that could be exploited to further sensitize resistant tumors or select the best trial candidates in future studies. Despite extensive multiregion sampling, we identified relatively few genes that were significantly altered in solely one group; all cases demonstrated hallmarks of aggressive tumors. However, amongst exceptional responders, which we defined as harboring residual tumor volumes < 0.05 cc, these somatic alterations were clonal, and histologic phenotypes were homogeneous. In contrast, tumors that demonstrated therapeutic resistance had greater subclonal heterogeneity, and phylogenetic reconstruction indicated more complex and branched evolutionary paths, such that both genomic and histologic diversity at baseline positively correlated with residual tumor.

The long-recognized variability and heterogeneity of localized prostate cancer has demonstrated that branching of the tumor occurs at key times during its development, including upon the transition from high-grade PIN to cancer, from low-grade to higher-grade cancers, and from localized disease to metastasis ^4,22,37,38^. Although these earlier studies demonstrated selection bias in the analysis of larger tumors that were more conducive to multiregion sampling and sequencing, more recent cohort-based molecular analyses have demonstrated strong relationships between tumor evolution and aggressivity ^5,6^. In the present study, we attempt to subdivide otherwise high-risk tumors based on their evolutionary state at the time of treatment. The finding that larger tumors were more likely to resist therapy^18^ fit well with the model that they had undergone more cell divisions since originating, with more opportunities for diversity and punctuated evolutionary events. Meanwhile smaller and more homogenous tumors were thus more sensitive, despite harboring features of advanced cancers such as higher Gleason grades and cribriform glandular architecture. Importantly, even smaller tumors in our study were sampled comprehensively, such that our bioinformatic reconstruction of tumor phylogenies was not biased by less complex data in ER cases. By definition, an ER case meant that all MRI-visible lesions responded exceptionally well. At the patient level, a subset of INR tumors demonstrated objective heterogeneity because they included lesions that responded as well as lesions that persisted ^18^, with the intrinsic variability associated with intrapatient response characteristics. At the extreme end of this phenomenon was a patient in our study who harbored two completely independent tumors that exhibited differential responses to therapy ^20^.

By comparing the histologic intensity of IHC in this cohort of patients to pathologic outcome, we found that PSA was inversely proportional, while GR was directly proportional. The fact that PSA levels tended to be uniformly high yet were heterogeneously reduced in INR cases is consistent with the hypothesis that AR-high tumors are more likely to respond to AR-directed therapy ^39,40^, and that lower PSA staining is representative of an AR-low subpopulation. Indeed, *in situ* PSA levels were reduced by treatment ^18^, with poor responders *a priori* less dependent upon AR. This finding is further supported by the observation that baseline GR histologic intensity increased with RCB post-treatment. Nuclear GR expressed in prostate cancer luminal cells is lower than in normal epithelium but is elevated in CRPC as one of several bypass mechanisms to reconstitute AR activity in the absence of bound ligand ^36,41^. In contrast to PSA, GR levels are uniformly low in our cohort but display heterogeneity in INR cases, potentially priming those tumors for resistance to inADT. Interestingly, with both stains, heterogeneity increased proportionally with RCB, and SDI’s calculated based on subpopulations of stained tumor cells also indicated greater *in situ* diversity. Thus by per-patient analysis, these direct phenotypes of prostate cancer offered visual and microscopic interpretation of subclonal diversity.

Genomically, our approach to sample tumors comprehensively also increased our sensitivity to identify mutations contributing to exceptional response. At the patient level, mutations exclusively found in ER cases included alterations to *SPOP*, which have been previously associated with response to ADT, are found with less frequency in mCRPC patients, and indicate a more favorable prognosis by conventional therapies ^42,43^. We also identified a limited number of genes with two-copy losses significantly enriched in ER cases. These include the proto-oncogene *FYN*, raising the fascinating possibility that alterations which contribute to tumorigenesis may sensitize the tumor to inADT, similar to the protective effects of null oncogene mutations ^44^. Identifying ER lesions in INR patients increased our ability to discover somatic alterations that may sensitize tumors to inADT. These lesion-level analyses also identified in ER lesions recurrent deletions to chromosome 5q14, which harbors *NR2F1*, a nuclear receptor associated with tumor cell dormancy ^45^; tumors with functioning NR2F1 may potentially evade inADT while deletion of that locus is sensitizing. Further *in vitro* and *in vivo* functional validation of these findings will shed light on mechanisms of exceptional response.

Our analysis also identified recurrent alterations that were enriched in INR patients. These events, which include the TMPRSS2:ERG fusion and loss of PTEN, are common in prostate cancer and have been observed more frequently in inADT-resistant tumors ^14,17^. We selected the histologic feature intraductal carcinoma (IDC-P) for use in our four-factor model also based on its strong enrichment in INR tumors. Recently, IDC-P has become increasingly appreciated for its association with poor overall outcome ^46,47^, and deeper investigations demonstrated relationships between IDC-P, hypoxia, and genomic instability ^48,49^. Indeed, PGA was also proportionally increased with both genomic SDI and the volume of treatment-resistant tumors. Incidentally, IDC-P was the only feature distinctly subclonal to INR cases, whereas chromosome 10q loss, TMPRSS2:ERG fusion and *TP53* mutations were generally clonal and thus happened early in the tumor’s history, explaining their increased incidence in ER samples by lesion-level analyses. Comparing the associations of each factor in our model to genomic SDI and prediction of INR outcome using a *t* test, IDC-P was the second most important feature (uncorrected *P* = 0.088), with only ERG showing the greatest importance (uncorrected *P* = 0.25). When *TP53* was omitted from our logistic regression model, ERG remained tightly associated with INR probability but no longer carried associations with genomic diversity, demonstrating the importance of the coincidence of these mutations for driving evolutionary processes. An important limitation of our approach in validating these factors is that until future studies with similarly comprehensive analyses of baseline tissue are performed, we will be unable to directly validate the outcome of residual tumor following inADT. However, prospective assessment of patients lacking these factors despite harboring high-risk prostate cancer may select for a greater proportion of ER cases in future studies.

Although inADT trials have been conducted over the past decade, the long interval between standard-of-care therapy, recurrence, and cancer-specific mortality has made it challenging to determine the precise overall survival benefits of inADT ^50,51^. However, if AR-targeted therapies active in metastatic hormone sensitive prostate cancer are reducing intraprostatic tumor volumes in a subset of patients, a reasonable hypothesis is that these agents also act on occult micrometastatic disease that might otherwise progress later. Indeed, in a meta-analysis of three inADT trials ^12,13,15,52^, patients who exhibited MRD or pCR (defined as < 0.5 cm residual tumor in the largest cross-sectional dimension) did not experience biochemical recurrence (BCR). However, in contrast to previous studies that had substantial numbers of patients with favorable-intermediate risk, our current study was enriched for unfavorable-intermediate and high-risk patients with greater chances of recurrence. Nonetheless, inADT would be expected to delay or prevent recurrence in at least a subset of the ER patients. Using the high-risk prostate TCGA cohort for which follow-up data was available, we compared the BCR-times by Kaplan-Meier analysis based on their predicted ER or INR status and did not observe a significant difference (Supplementary Fig. 8). This finding suggests that amongst higher risk patients, the four factors used in our model did not independently affect BCR, and by extension, that inADT had the potential to delay BCR in a subset of those patients.

Outcomes of patients with intermediate-to-high risk prostate cancer remain variable, but in the context of prior studies ^5,22,38^, connecting short-term responses to treatment with extensive histogenomic analyses has revealed that the evolutionary history of a tumor is clearly related to its fitness. Those patients with smaller tumors and more linear evolution paths demonstrated responses to earlier (preoperative) inADT, which is expected to delay or prevent recurrence. In contrast, those patients with more evolved tumor systems may harbor oligometastatic disease, which in conjunction with sensitive molecular imaging modalities such as PSMA positron emission tomography, could provide opportunities for radiation therapy and overall intensification of treatment. A study using inADT with imaging to evaluate intraprostatic tumor responses at multiple time points is currently in progress (ClinicalTrials.gov identifier: NCT02430480).

## Data Availability

Raw sequencing data has been deposited in the Database of Genotypes and Phenotypes (dbGaP) accession number phs001938.v2.p1.

## ACKNOWLEDGMENTS

The authors gratefully acknowledge the patients and the families of patients who contributed to this study. The authors thank Dr. David Takeda for his critical review of the manuscript. DNA sequencing was performed at the CCR Illumina Sequencing Facility and the CCR Genomics Technology Laboratory. Portions of this work utilized the computational resources of the NIH HPC Biowulf cluster.

## DATA AVAILABILITY STATEMENT

Raw sequencing data depicted in Figure 1 has been deposited in the Database of Genotypes and Phenotypes (dbGaP) accession number phs001938.v2.p1. Custom code for processing data has been made available at https://github.com/CBIIT/lgcp.

